# Impact of diabetes on longevity and disability-free life expectancy among older South African adults: a prospective longitudinal analysis

**DOI:** 10.1101/2022.07.31.22278253

**Authors:** Collin F. Payne, Lilipramawanty K. Liwin, Alisha N. Wade, Brian Houle, Jacques D. Du Toit, David Flood, Jennifer Manne-Goehler

## Abstract

**Objective:** We seek to understand the coexisting effects of population aging and a rising burden of diabetes on healthy longevity in South Africa.

**Research Design and Methods:** We used longitudinal data from the 2015 and 2018 waves of the “Health and Aging in Africa: A Longitudinal Study of an INDEPTH Community in South Africa” (HAALSI) study to explore life expectancy (LE) and disability-free life expectancy (DFLE) of adults aged 45 and older with and without diabetes in rural South Africa. We estimated LE and DFLE by diabetes status using Markov-based microsimulation.

**Results:** We find a clear gradient in remaining LE and DFLE based on diabetes status. At age 45, a man without diabetes could expect to live 7.4 [95% CI 3.4 – 11.7] more years than a man with diabetes, and a woman without diabetes could expect to live 3.9 [95% CI: 0.8 – 6.9] more years than a woman with diabetes. Individuals with diabetes lived proportionately more years subject to disability than individuals without diabetes. Additional analyses separating individuals with diabetes based on whether they knew their disease status found that individuals with diabetes diagnosed by a healthcare worker had shorter remaining LE than those who were unaware of their status or those without diabetes.

**Conclusions:** Our findings highlight the large and important decrements in healthy, disability-free aging for people with diabetes in South Africa. This finding should motivate efforts to strengthen prevention and treatment efforts for diabetes and its complications for older adults in this setting.

---

Diabetes is an urgent global health problem that in 2019 was ranked as the third leading risk factor for morbidity and mortality worldwide. (1) The diabetes epidemic also has become increasingly well-recognized as a growing public health crisis in sub-Saharan Africa, where 24 million people are currently living with diabetes, including over 4 million in South Africa alone. Current projections suggest that – without intervention – more than 55 million people will be living with diabetes in the region by 2045. (2) In addition to this exploding population-level burden of diabetes, there is compelling evidence that health systems in South Africa and throughout the region have struggled to keep up with the need for diagnosis, treatment, and control of diabetes, with few adults with diabetes achieving comprehensive risk factor control in this setting. (3–5)

The prevalence of diabetes tends to increase as adults enter middle age, and, in studies from high-income countries, diabetes confers an increased risk of morbidity and mortality among older adults. (6) However, the relationship between diabetes and important aging-related outcomes in resource-limited settings remains poorly understood. Several large cohort studies have reported that people with diabetes in Asia and Latin America have 2-4 times greater mortality rates than people without diabetes. (7–9) In Sub-Saharan Africa, population-based data on diabetes outcomes are extremely limited, though the Prospective Urban Rural Epidemiologic (PURE) study recently reported that diabetes was associated with an age- and sex-adjusted mortality hazard ratio of 1.60 across sites in South Africa, Tanzania and Zimbabwe. (10) While these studies have begun to clarify the impact diabetes can have on mortality, there is still a large gap in understanding how diabetes influences healthy longevity outside of high-income contexts. These relationships are of particular interest in sub-Saharan African settings where communicable disease epidemics such as HIV have been dominant and health systems capacity for chronic diseases such as diabetes has been limited. (11)

In this study, we aim to evaluate the relationship between diabetes and healthy longevity among older adults in South Africa. To do so, we take advantage of the population-based “Health and Aging in Africa: A Longitudinal Study of an INDEPTH Community in South Africa” (HAALSI) longitudinal cohort, which collected data from 2015 to 2018. Using Markov-based microsimulation, we estimate both life expectancy (LE) and disability-free life expectancy (DFLE) among people with and without diabetes, and further explore differences in both outcomes for people who carry a formal diabetes diagnosis as compared to those who do not.

## Research Design and Methods

### Study design and Data

This was an observational study using Markov-based microsimulation models to estimate LE and DFLE among individuals with and without diabetes in the HAALSI longitudinal cohort.HAALSI is a Health and Retirement sister study based in the Agincourt health and demographic surveillance site in Mpumalanga, South Africa, with waves of data collected in 2015 and 2018. (12) As compared to most studies of older adults, the HAALSI cohort uses a lower starting age (age 40), as a key study objective is to understand the risk of incident cognitive impairment and its relationship to both HIV and cardiometabolic disease risk factors in aging adults. Details of the HAALSI cohort, survey scope, data collection, and sampling frame are available in a Cohort Profile.(13) Ethical approval for HAALSI was obtained from the University of the Witwatersrand Human Research Ethics Committee (no. M141159), the Harvard T.H. Chan School of Public Health Office of Human Research Administration (no. 13–1608), and the Mpumalanga Provincial Research and Ethics Committee. HAALSI survey data are publicly available at https://dataverse.harvard.edu/dataverse/haalsi.

### Measures

Our primary measure of functional health is based on the Activities of Daily Living scale (14,15). We generated two distinct states of physical health: **disability-free** individuals with no reported limitations on ADL activities, and **ADL disabled** individuals with one or more limitation on ADL activities. ADL disability is defined as reporting difficulty on any of the following six activities: dressing, bathing, eating, getting in/out of bed, toileting, and walking across a room. Where necessary, proxy responses on ADL limitation were used (N=84 in 2015, N=208 in 2018).

In the 2015 data collection, anthropometry, blood pressure, point-of-care glucose and dried blood spots for HIV testing were collected. Participants provided consent for participation in the survey and specimen collection. Participants were not instructed to fast. Fasting status was determined based on the timing of the last reported meal. Diabetes was defined as a fasting capillary glucose (defined as time since last meal > 8 hours) of ≥7.0 mmol/L or random capillary glucose ≥11.1 mmol/L measured at the time of baseline interview using a CareSens N blood glucose meter, or self-reported previous diagnosis of diabetes mellitus by a doctor, nurse or healthcare worker (16,17). We additionally explored how healthy longevity varied depending on prior awareness of individuals’ diabetes diagnosis. Individuals who reported a prior diagnosis of diabetes are classified as having **diagnosed diabetes**, and those who recorded a blood glucose measurement that met criteria for diabetes during the survey but had not been previously diagnosed are classified as having **undiagnosed diabetes**.

A total of 5,059 eligible men and women aged 40 years or over consented to participate and were included in the HAALSI survey in 2015 (85.9% response rate). Of those participating, 4,652 respondents provided a sample for point-of-care glucose testing and returned a valid result, and 4,640 of these provided responses to the ADL questions. In 2018, 3,836 (83%) of these individuals were re-interviewed, 535 (12%) had died between survey waves, 231 (5%) refused to be re-interviewed, and 38 (<1%) could not be found. Incident mortality of individuals in the HAALSI dataset is tracked by the Agincourt Health and Demographic Surveillance System.

### Statistical methods

Our primary outcome measures are total life expectancy (LE) and disability-free life expectancy (DFLE), compared across the populations with and without diabetes. LE and DFLE are metrics that compare how longevity and physical functioning differ across population subgroups. DFLE divides life expectancy into life-years spent free of ADL limitations and years spent subject to an ADL disability, combining mortality and health into a comprehensive metric for measuring healthy longevity at the population level.

We estimated LE and DFLE by diabetes status using microsimulation-based multistate life tables. (18) We modeled the annual probabilities of transitioning between disability states (described in Supplemental Figure 1) using a cumulative logistic regression model, stratified by initial disability state. The model includes age and age2 as continuous predictors; sex and diabetes status (diabetes vs no diabetes) as categorical variables; several interaction terms: age * diabetes status, age * sex, and sex * diabetes status; and relaxes the proportional odds assumption on sex and diabetes status.(19) An attrition weight was generated to account for nonresponse due to reasons other than mortality between waves 1 and 2 (e.g., refusal or not found for contact). This weight generating model included basic sociodemographics (age, sex, birthplace, level of schooling, marital status), migration history, and information on contact with the survey team.(20) This final attrition weight was included in the model estimating the transition probabilities.(21) We then predicted matrices of age-specific transition probabilities for each combination of sex and diabetes status, separately by initial disability state. Graphs of these annual transition probabilities are provided in Supplemental Materials.

These observed transition probabilities were then used as the basis for a microsimulation-based multistate life table model, using an adapted version of the SPACE suite of SAS programs (18,22). We generated synthetic cohorts of 100,000 individuals at initial ages of 45 and 65. These synthetic cohorts were assigned the same sex, diabetes status, and disability state distribution as observed in the HAALSI data: the average characteristics of the HAALSI cohort aged 40-49 were used to generate the synthetic cohort at age 45, and the average characteristics of the HAALSI cohort aged 60-69 were used to generate the synthetic cohort at age 65. We then “aged” these individuals forward year by year via microsimulation, using the age-, gender-, and diabetes-status-specific mortality rates and probabilities of transitioning in and out of disability estimated above. The resulting synthetic cohort, representing the simulated life courses of 100,000 individuals subjected to the transition rates observed in the HAALSI data between 2015 and 2018, was analyzed to estimate total LE and DFLE. Point estimates shown were from the transition probabilities estimated from the full sample. Confidence intervals (CIs), which reflect both the uncertainty of the estimated parameters and the uncertainty from the microsimulation, were created by re-estimating the above analysis sequence using 499 bootstrap re-samples for each age group under study. We took the central 95% of the distribution of these 500 parameters (the estimates from the full sample, and 499 bootstrap resamples) as the 95% confidence interval.

### Role of the funding source

The funder of the study had no role in study design, data collection, data analysis, data interpretation, or writing of the report.

## Results

### Baseline characteristics

Baseline characteristics of the analysis sample by diabetes status are provided in Table 1. Overall, 89% of the sample were classified as not having diabetes, and 11% were classified as having diabetes by biomarker criteria or prior diagnosis. The sample with diabetes was on average slightly older than population without diabetes. The HAALSI sample has more women than men, a result of sex differences in both survivorship and labor migration, and women were also slightly overrepresented among the sample with and without diabetes.

**Table 1:** Sample characteristics and age-standardized health indicators, by diabetes.

| **Characteristic** | **No**<br>**diabetes**<br>**(%)** | **Diabetes**<br>**(%)** | **Diagnosed**<br>**diabetes**<br>**(%)** | **Undiagnosed**<br>**diabetes**<br>**(%)** |
| --- | --- | --- | --- | --- |
| **N** | (4082) | (557) | (336) | (221) |
| % | 89.1 | 10.9 | 60.3 | 39.7 |
| **Age** |  |  |  |  |
| 40-49 | 94.3 | 5.7 | 44.4 | 55.6 |
| 50-59 | 89.2 | 10.8 | 60.5 | 39.5 |
| 60-69 | 86.1 | 13.9 | 59.7 | 40.3 |
| 70+ | 85.0 | 15.0 | 64.1 | 35.9 |
| **Sex** |  |  |  |  |
| Male | 46.9 | 41.5 | 61.7 | 38.3 |
| Female | 53.1 | 58.5 | 59.3 | 40.7 |
| **Health indicators (age standardized)** |  |  |  |  |
| **ADL Disability** |  |  |  |  |
| No ADL limitations | 92.1 | 85.5 | 82.2 | 91.0 |
| 1+ ADL limitation | 7.9 | 14.5 | 17.8 | 9.0 |
| **Co-morbidities** |  |  |  |  |
| **Hypertension<sup>a</sup>** |  |  |  |  |
| Yes | 61.7 | 79.0 | 87.1 | 66.4 |
| No | 38.3 | 21.0 | 12.9 | 33.6 |
| **Dyslipidemia<sup>b</sup>** |  |  |  |  |
| Yes | 41.6 | 60.4 | 60.1 | 60.7 |
| No | 58.4 | 39.6 | 39.9 | 39.3 |
| **HIV** |  |  |  |  |
| HIV Negative | 76.1 | 84.7 | 85.0 | 84.3 |
| HIV Positive | 23.9 | 15.3 | 15.0 | 15.7 |
| **BMI classification<sup>c</sup>** |  |  |  |  |
| Underweight | 5.6 | 1.9 | 1.3 | 6.5 |
| Normal | 38.5 | 23.4 | 23.1 | 38.6 |
| Overweight | 28.1 | 29.3 | 31.1 | 28.9 |
| Obese | 27.5 | 45.2 | 44.5 | 26.0 |

Notes: ADL=activities of daily living; BMI = body mass index.
Data were missing for 17 individuals on hypertension, 399 individuals on dyslipidemia, 139 individuals on HIV status, and 81 individuals on body mass index.
<sup>a</sup> Hypertension defined as Systolic BP ≥ 140 mmHg or diastolic BP ≥ 90 mmHg, or reporting using anti-hypertensive medication at time of interview
<sup>b</sup> Dyslipidemia is defined as elevated total cholesterol ( ≥ 6.21 mmol/L) or low HDL (1.19 mmol/L) or elevated LDL ( $>$ 4.1 mmol/L) or elevated triglycerides ( $>$ 2.25 mmol/L) or self-report of ever diagnosed with high cholesterol
<sup>c</sup> BMI classification used the following cutoffs: BMI $<$ 18.5 = underweight; BMI 18.5-24.9 = Normal; BMI 25-29.9 = overweight, BMI ≥ 30 = obese.

Participants were approximately evenly divided across the four age categories. After age-standardizing to remove the effect of age-compositional differences between individuals with and without diabetes, a total of 8.5% of the sample without diabetes and 15% of sample with diabetes had at least one as ADL disability. Individuals with diabetes were substantially more likely to have hypertension and dyslipidemia than individuals without diabetes. Over 45% of individuals with diabetes were classified as obese, defined as BMI ≥30 kg/m2, as compared to just over one quarter of the sample without diabetes. However, individuals with diabetes were less likely to be HIV positive, 14.3% vs 22.4%. Among individuals with diabetes, about 60% had received a diagnosis by a healthcare worker. Individuals with diagnosed diabetes were on average older, more likely to have an ADL disability, more likely to be obese, and more likely to be hypertensive than individuals with undiagnosed diabetes.

### Health transition probabilities

As a first step to estimating LE and DFLE by diabetes status, we modeled the underlying age-, sex-, and diabetes-status-specific annual transition probabilities between disability-free and ADL disabled life. Panel A of Figure 2 displays the annual transition probabilities out of disability-free life by sex and diabetes status, along with 95% CIs based on 499 bootstrap resamples. As expected, transition probabilities from disability-free life to ADL disabled life and death rose substantially with age, and we found that transitions from disability-free life to ADL disabled life and to death were higher among the population with diabetes than without diabetes. The likelihood of transitioning to ADL disabled life was substantially lower among men than among women, and annual probabilities of mortality among men with diabetes rose rapidly over age. With increasing age, ADL disabled individuals were substantially less likely to transition back to disability-free life (Panel B of Figure 2), and recovery from ADL disability was much less likely among individuals with diabetes than those without diabetes. By age 63, an ADL disabled man with diabetes had a higher likelihood of dying over the next year than of recovering to disability-free life (the corresponding age for a man without diabetes was 77). At every age, ADL disabled individuals with diabetes were more likely to die than their counterparts who did not have diabetes.

**Figure 2:**
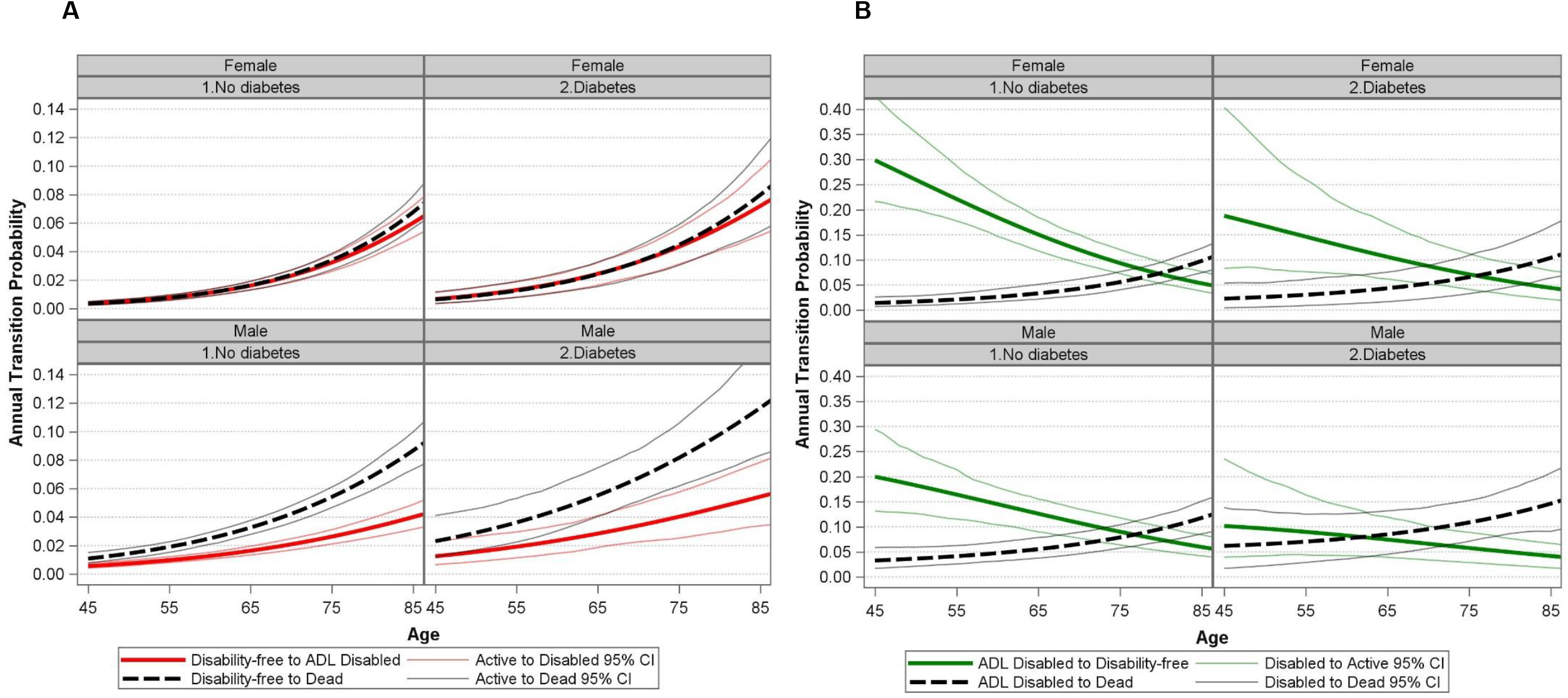
Transition probabilities from disability-free (Panel A) and ADL disabled (Panel B) life by age and diabetes status

### Life and disability-free life expectancies by diabetes status

Figure 3 and Supplemental Table 1 present LE and DFLE for men and women in the HAALSI cohort at ages 45 and 65 by diabetes status. At age 45, a man without diabetes could expect to live an additional 26.8 [95% CI: 25.2 - 28.4] years, over seven years longer than the life expectancy of a man with diabetes, which was 19.4 [95% CI: 15.6 - 22.8] years. Remaining life expectancies were higher for women, but similar trends were seen—LE at age 45 was 33.7 [95% CI: 32.4 - 35.1] among women without diabetes, nearly four years longer than for women with diabetes (29.8 [95% CI: 26.3 - 33.2]). These gaps in remaining LE were smaller but still substantial at older ages: at age 65, a man who did not have diabetes could expect to live another 14.5 [95% CI: 13.8 - 15.5] years, over three years more than the 11.4 [95% CI: 9.6 - 12.8] year LE of a man with diabetes (for women, these figures are 17.5 [95% CI: 16.8 - 18.5] years among women without diabetes and 15.7 [95% CI: 13.9 - 17.7] years among women with diabetes).

**Figure 3:**
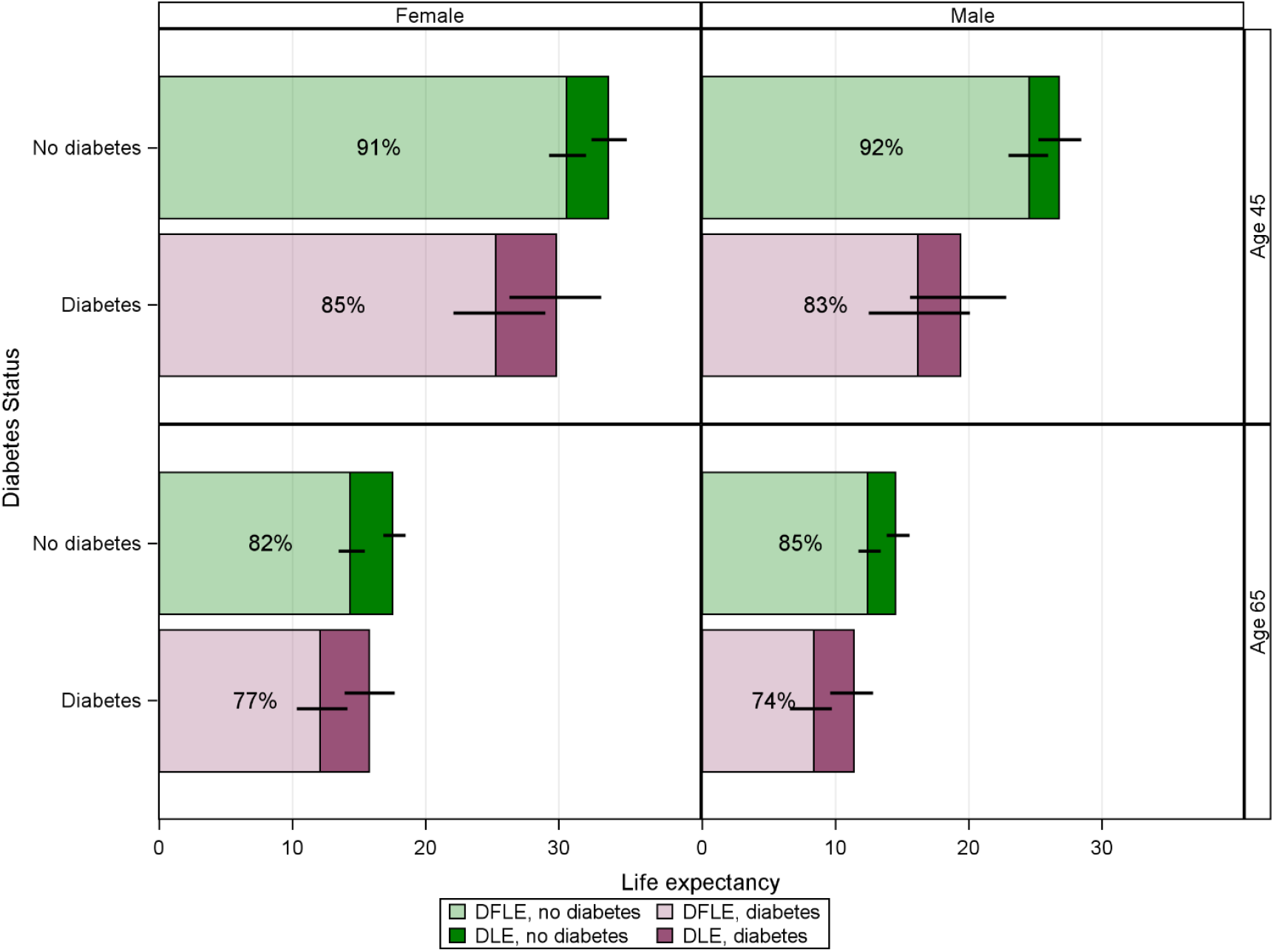
Microsimulation estimated disability-free, disabled, and total remaining life expectancy at ages 45 and 65 by diabetes status, HAALSI 2015-2018 **Notes**: Black bars represent the 95% confidence interval around point estimates of LE and DFLE. Percentage figures refer to the percent of total LE spent disability-free. DFLE = disability-free life expectancy, DLE = ADL disabled life expectancy.

Turning to DFLE, we find that individuals with diabetes in this rural South African population spend substantially more of their remaining life subject to limitations on ADL activities. At age 45, a man with diabetes could expect to spend only an additional 16.1 [95% CI: 12.5 - 20.1] years of life free of ADL disability, compared with 24.5 [95% CI: 23.0 - 25.9] disability-free years for a man with no diabetes. These figures translate to a man with diabetes spending 83% [95% CI: 0.75 - 0.92] of his already shorter remaining LE free of ADL limitations, as compared to 92% [95% CI: 0.89 - 0.94] for a man without diabetes.Differences in DFLE between women with and without diabetes women were slightly smaller, but at age 45 a woman without diabetes could expect to spend an additional 30.6 [95% CI: 29.3 - 32.0] years free of ADL limitations, over 5 more disability-free years than for a woman with diabetes (25.3 [95% CI: 22.1 - 29.0] years). Figures were overall similar at age 65, with substantial gaps in DFLE between individuals with and without diabetes.

### Life and disability-free life expectancies by self-knowledge of diabetes status

We additionally sought to understand how awareness of diabetes status may impact healthy longevity. This stratification was important because awareness of diabetes diagnosis may indicate care-seeking and engagement in lifestyle modification or treatment activities that could in turn impact the trajectory of diabetes, its complications and risk of disability. In these analyses, we substituted a three-category measure of diabetes status (no diabetes, diagnosed diabetes, and undiagnosed diabetes) in place of the binary diabetes status described in the methods section above.

Figure 4 and Supplemental Table 2 provide the results of these analyses. Among both men and women, LE estimates for those with diagnosed diabetes trailed those of both the undiagnosed diabetes and no diabetes groups. For a woman at age 45, an individual with diagnosed diabetes could expect to live 27.7 (95% CI 23.8-31.5) years, over six years fewer than a woman with no diabetes (33.8 years [95% CI 32.6 - 35.5]); a woman with undiagnosed diabetes had only a 1.5 year shorter life expectancy (32.3 years [95% CI 27.2 - 36.5]). For both men and women, individuals with diagnosed diabetes could expect to live more of their remaining years disabled, a pattern that held at age 45 and at age 65. Given the small sample sizes in the groups with diagnosed, undiagnosed, and no diabetes, confidence intervals around our estimates are substantial and differences between groups are not universally significant. Of note, the gradient in LE and DFLE between these groups was quite similar across gender and initial age.

**Figure 4:**
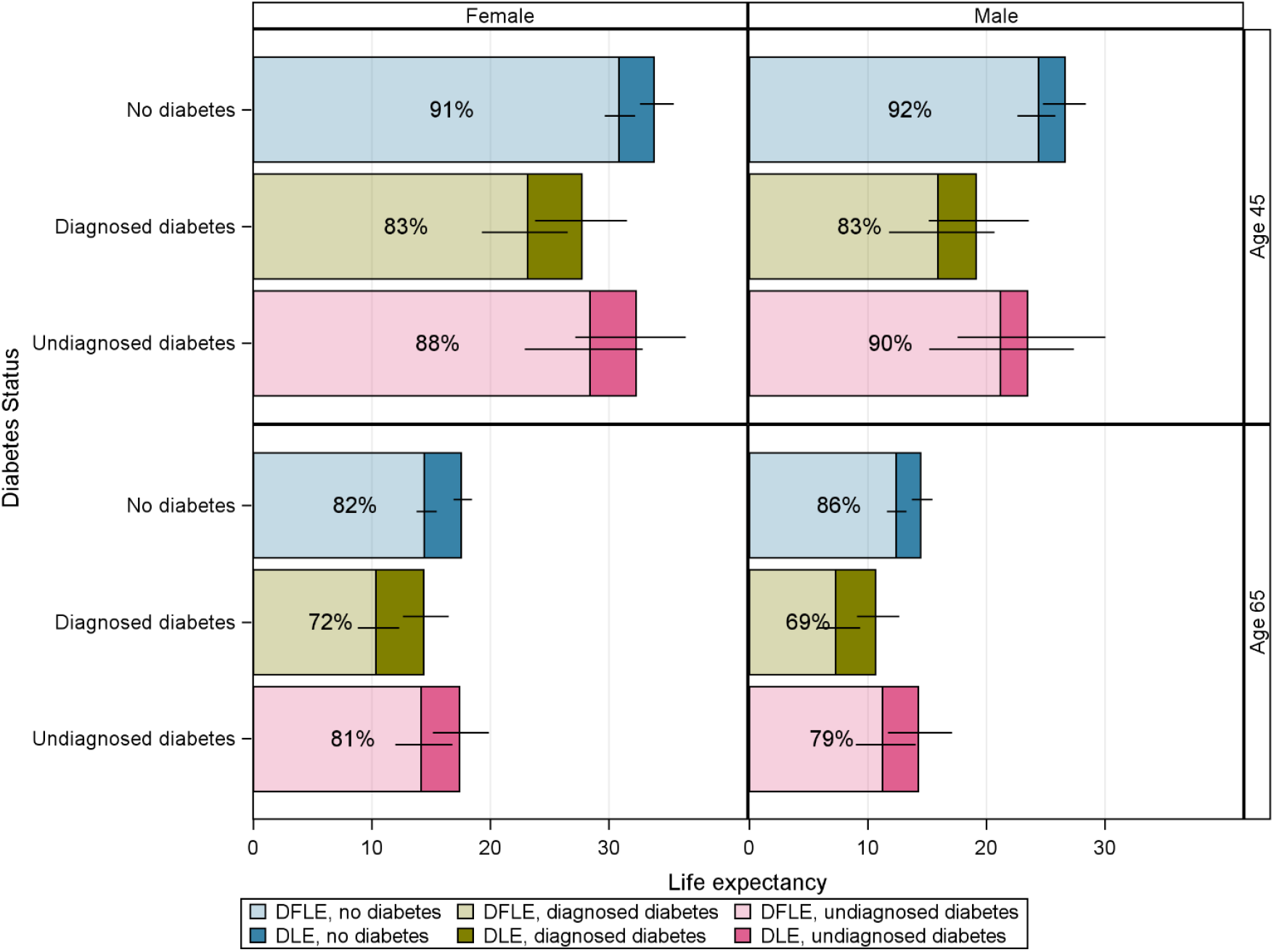
Microsimulation estimated disability-free, disabled, and total remaining life expectancy at ages 45 and 65 by diabetes status and knowledge of status, HAALSI 2015-2018 **Notes**: Black bars represent the 95% confidence interval around point estimates of LE and DFLE. Percentage figures refer to the percent of total LE spent disability-free. DFLE = disability-free life expectancy, DLE = ADL disabled life expectancy.

## Conclusions

In this study using longitudinal data from a population-based cohort in South Africa collected between 2015 and 2018, we found that people with diabetes experienced substantially lower life expectancy and spent a greater proportion of their lives with a disability compared to people without diabetes. At age 45 years, a man with diabetes lived 7-8 years less than a man without diabetes and spent 1 additional year living with a disability while a woman with diabetes lived 3-4 years less than a woman without diabetes and spent 1-2 years additional years living with a disability. To our knowledge, this study is the first to investigate how diabetes shapes healthy longevity in Sub-Saharan Africa and one of the first studies of its kind conducted outside of a high-income country.

Much of the existing literature on the population-impact of diabetes in low- and-middle income countries focuses on the relationship between diabetes and mortality; few studies have examined how diabetes shapes other important aging-related outcomes such as physical disability. The only prior study we have identified that assesses total and disability-free life expectancy among people with diabetes in a low or middle-income country used data from the 2001 to 2003 waves of the Mexican Health and Aging Study (23). This study found that, at 50 years of age, diabetes was associated with approximately 10 years less total life expectancy and 9 years less disability-free life expectancy (defined using ADLs). In high-income countries, several studies have shed insight on the disability-adjusted life expectancies of people with versus without diabetes. For instance, a study using data collected between 1998 to 2012 from the U.S. Health and Retirement Study found that people with diabetes at age 50 years, compared to people without diabetes, could expect to have a shorter life expectancy by approximately 4-5 years and spend 1-2 additional years living with an ADL-defined disability. Similar results have been reported in other high-income countries such as Canada and Australia (24,25). While direct comparisons between our study and studies conducted in other countries are challenging due to temporal and methodological differences, our findings suggest that diabetes in our cohort in South Africa is associated with a larger decrement in both total life expectancy and disability-free life expectancy than in high-income contexts. One driver of this finding may be the lack of preventive care or therapeutic support for common diabetes complications including retinopathy, neuropathy, cardiovascular complications and ulcers.(11)

Our finding that women with diabetes live longer but experience a greater number of years with a disability is an important secondary finding that has also been observed in prior studies (23,24,26). Nevertheless, the disparities observed in our South African data are striking and suggest that diabetes is associated with a substantially more negative impact on men than women. Globally, men with diabetes are less likely than women to receive diabetes treatment recommended in clinical guidelines (3). In South Africa, prior studies suggest that women are more likely than men to receive some health care services for noncommunicable diseases and to achieve glycemic control (27,28). Taken together, these findings imply a need to better understand the underlying mechanisms leading to gender-based differences in diabetes outcomes and to develop policies to mitigate these disparities.

These findings have implications for both policy responses to the growing diabetes epidemic and as a basis for future research into the relationship between diabetes and important aging outcomes. The trends of growing diabetes prevalence and its impact on healthy life expectancy in South Africa suggest large potential benefits in strengthening systems of care for this disease to promote healthy aging. First, diabetes is a potentially preventable condition, but few resources have been expended to prevent or delay the onset of the disease at the population level in this region.(11) Additionally, prior research has shown that many adults with diabetes are unable to achieve control of their disease, as well as other comorbid risk factors. (28,29) Efforts to improve health system performance for diabetes may be motivated by a greater understanding of the toll diabetes takes on both life expectancy and healthy longevity. Second, evidence from high-income settings has suggested that there may be more than one subtype of diabetes among aging adults and that management strategies may need to be tailored to maximize healthy aging in these distinct sub-groups. (30) Third, more research is needed to identify the complications that lead to loss of healthy life expectancy in people with diabetes. For instance, common complications of diabetes such as loss of eyesight and neuropathy can affect aging in important ways but have not been a major focus of existing policies or programs in the region to date.

A strength of our study compared to the similar analysis conducted in Mexico was that we did not rely solely on self-report to define diabetes exposure. Rather, we used a blood-based diabetes biomarker, glucose, to define diabetes status among people without a history of previously diagnosed diabetes. When we separated people with diabetes into diagnosed versus undiagnosed categories, it appeared as though individuals with diagnosed diabetes had lower LE and DFLE compared to individuals with undiagnosed diabetes, although small samples sizes precluded precise confidence intervals for these estimates. These findings are broadly consistent with global literature showing greater mortality among people with self-reported diabetes relative to undiagnosed diabetes (7,10). It is likely that people with diagnosed diabetes represent a distinct subpopulation with greater diabetes severity and duration than people with undiagnosed diabetes, though, at the same time, this population may also have better access to health care services. While more data are needed on this topic, one policy implication emerging is that health system resources may be better invested in strengthening the quality of diabetes treatment and management of diabetes complications, rather than implementing widespread diabetes screening programs.

This study is subject to several limitations. First, our methodological approach does not allow us to control for confounding characteristics that are often clustered with diabetes such as obesity, hypertension, dyslipidemia, and other cardiometabolic risk factors. As such, our findings should be interpreted as the average difference between individuals with diabetes and individuals without diabetes (who may, on average, differ on multiple characteristics beyond their diabetes status) and not as the causal effect of diabetes. We justify this approach as reasonable to achieve our primary objective of estimating the total and disability-free life expectancy among people with diabetes versus those without diabetes. In future research, we plan to investigate the underlying causal mechanisms driving the risks of disability and death among diverse populations of older adults with diabetes around the world, including from the HAALSI cohort in South Africa analyzed in this study.

Second, individuals with diabetes in our sample had a substantially lower prevalence of HIV (14.3% vs. 22.4%) as compared to those without diabetes. Given that HIV is independently associated with reductions in healthy aging in the HAALSI cohort (31 IN PRESS), our findings may underestimate the extent to which diabetes is associated with decrements in healthy aging among our sample. Third, among people without a self-reported diabetes diagnosis, we defined diabetes status using a single capillary glucose measurement. While single glucose measurements are standard in population-based diabetes surveys (32), clinical guidelines require repeat testing to confirm diabetes diagnosis (33,34). Fourth, our analyses do not account for incident diabetes cases between 2015 and 2018, as we lack follow-up information on diabetes status for individuals who died between waves. Fifth, our definition of disability using ADLs represents a severe level of functional limitation. ADL deficits are clinically very significant and thus appropriate for our analysis, but we suggest caution in generalizing our results to other facets of health, as trends in lower-level functional limitations may not follow the same patterns of ADL disability. A final limitation of our methodological approach is that the multistate analyses follow a simple Markov logic and are not state-duration-dependent. Individuals who experience a disability-state transition between survey waves are assumed to experience only a single transition over this period, which likely misses shorter-term fluctuations in functional health including incident disability prior to death (35).

In conclusion, in one of the first studies quantifying healthy longevity among people with diabetes outside of a high-income context, we found that diabetes was associated with large reductions in life expectancy and disability-free life expectancy in South Africa. Given the rising prevalence of diabetes and rapid population aging in low- and middle-income countries and Sub-Saharan Africa, our findings show the urgent need to improve diabetes care globally and to better understand the mechanisms of healthy aging among diverse global populations with diabetes.

## Data Availability

Data from the HAALSI study are publicly available at https://dataverse.harvard.edu/dataverse/haalsi.

## Acknowledgments

This work was supported by the National Institute of Aging at the National Institutes of Health (grant numbers 1P01AG041710–01A1, HAALSI – Health and Aging in Africa: A Longitudinal Study of an INDEPTH Community in South Africa). The content is solely the responsibility of the authors and does not necessarily represent the official views of the National Institutes of Health. The HAALSI study is nested within the Agincourt Health and socio-Demographic Surveillance System, a node of the South African Population Research Infrastructure Network (SAPRIN) supported by the Department of Science and Innovation, the University of the Witwatersrand, and the Medical Research Council, South Africa, and previously the Wellcome Trust, UK (Grants 058893/Z/99/A; 069683/Z/02/Z; 085477/Z/08/Z; 085477/B/08/Z). CFP is supported by an Australian Research Council Discovery Early Career Researcher Award (project number DE210100087) funded by the Australian Government, and by an ANU Futures Scheme Award funded by the Australian National University. ANW is supported by the Fogarty International Centre, National Institutes of Health [grant number K43TW010698].

## Conflict of Interest

No potential conflicts of interest relevant to this article were reported.

## Contributors

CFP, DF, and JM-G conceived the study. CFP wrote the analytic plan with input from LKL, ANW, BH, JDDT, DF, and JM-G. CFP carried out the analysis, and wrote the first draft together with DF and JM-G. All authors critically revised the manuscript and approved the final version. CFP is the guarantor of this work and, as such, had full access to all the data in the study and takes responsibility for the integrity of the data and the accuracy of the data analysis. All authors had full access to all the data in the study and had final responsibility for the decision to submit for publication.

---

**Supplemental Figure 1:**
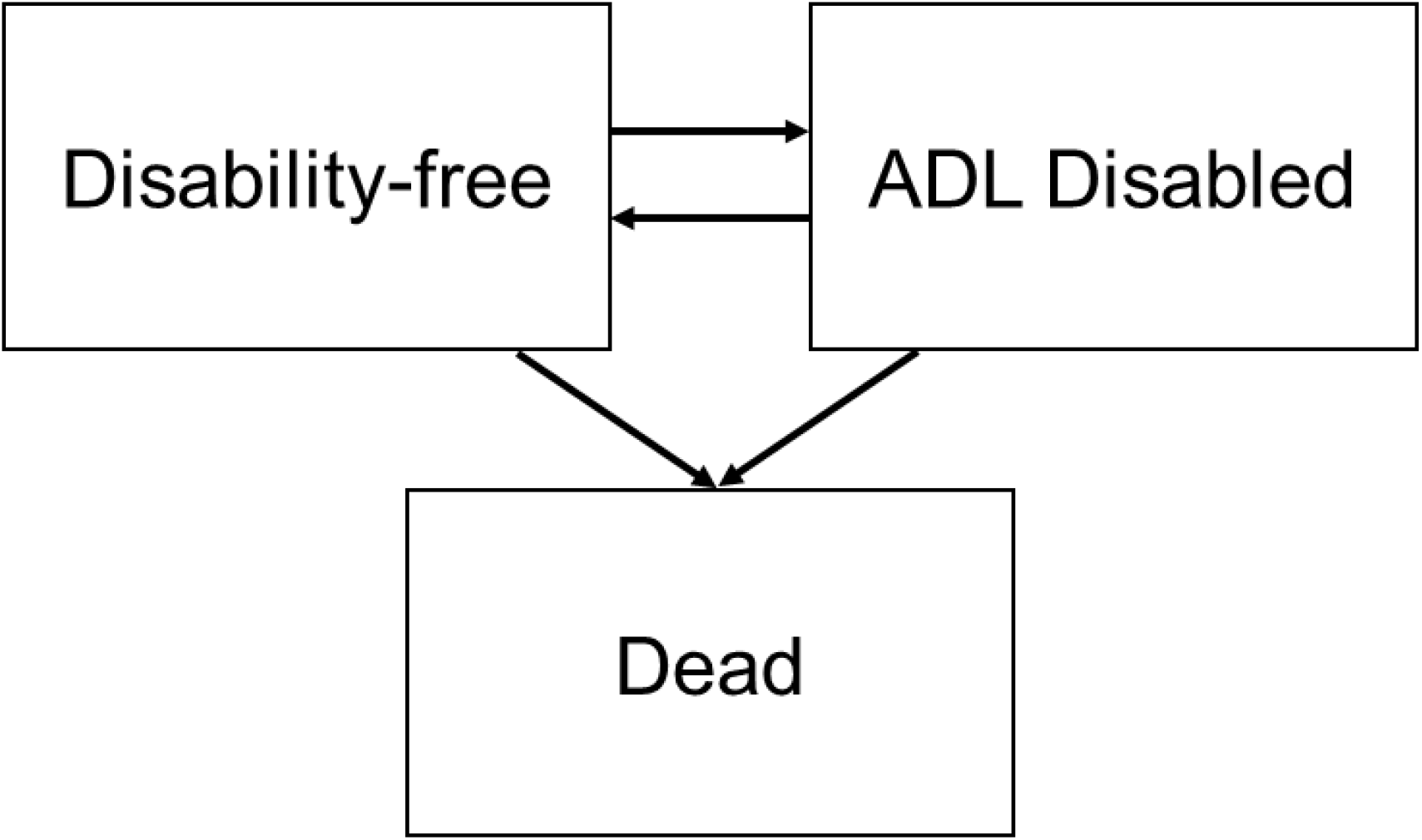
State-space for multistate model

**Supplemental Table 1:** Microsimulation estimated disability-free, disabled, and total remaining life expectancy (LE) at ages 45 and 65 by diabetes status, HAALSI 2015-2018

|  | Total LE | Disability-free LE | ADL disabled LE | Proportion disability-free |
| --- | --- | --- | --- | --- |
|  | Estimate [95% CI] | Estimate [95% CI] | Estimate [95% CI] | Estimate [95% CI] |
| **<u>Males</u>** |  |  |  |  |
| **Age 45** |  |  |  |  |
| No diabetes | 26.78 [25.22 - 28.44] | 24.54 [22.97 - 25.94] | 2.23 [1.69 - 2.81] | 0.92 [0.89 - 0.94] |
| Diabetes | 19.37 [15.59 - 22.79] | 16.14 [12.48 - 20.09] | 3.22 [1.64 - 4.93] | 0.83 [0.75 - 0.92] |
| **Age 65** |  |  |  |  |
| No diabetes | 14.51 [13.84 - 15.54] | 12.40 [11.70 - 13.39] | 2.11 [1.69 - 2.65] | 0.85 [0.82 - 0.88] |
| Diabetes | 11.38 [9.57 - 12.80] | 8.37 [6.55 - 9.73] | 3.01 [1.96 - 4.23] | 0.74 [0.63 - 0.82] |
| **<u>Females</u>** |  |  |  |  |
| **Age 45** |  |  |  |  |
| No diabetes | 33.69 [32.44 - 35.08] | 30.57 [29.27 - 32.04] | 3.13 [2.46 - 3.77] | 0.91 [0.89 - 0.93] |
| Diabetes | 29.80 [26.26 - 33.17] | 25.27 [22.07 - 28.95] | 4.53 [3.03 - 6.19] | 0.85 [0.80 - 0.90] |
| **Age 65** |  |  |  |  |
| No diabetes | 17.52 [16.82 - 18.51] | 14.33 [13.45 - 15.41] | 3.19 [2.60 - 3.85] | 0.82 [0.78 - 0.85] |
| Diabetes | 15.74 [13.91 - 17.67] | 12.07 [10.32 - 14.13] | 3.67 [2.63 - 4.78] | 0.77 [0.70 - 0.84] |

Notes: DFLE = disability-free life expectancy, DLE = ADL disabled life expectancy.

**Supplemental Table 2:**
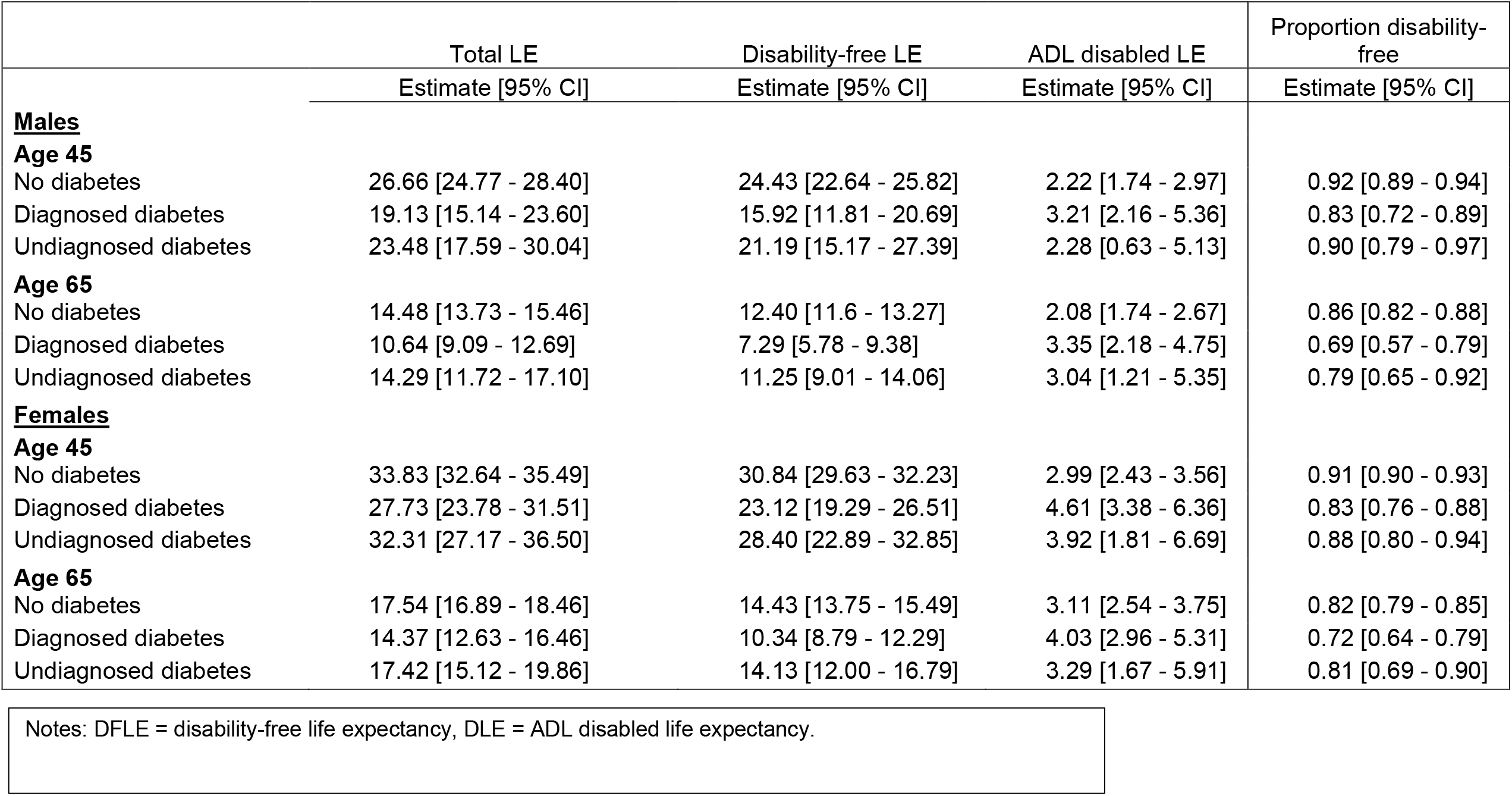
Microsimulation estimated disability-free, disabled, and total remaining life expectancy (LE) at ages 45 and 65 by diabetes status and self-knowledge, HAALSI 2015-2018

## Notes

### Competing Interest Statement

The authors have declared no competing interest.

### Author Declarations

Ethical approval for HAALSI was obtained from the University of the Witwatersrand Human Research Ethics Committee (no. M141159), the Harvard T.H. Chan School of Public Health Office of Human Research Administration (no. 13-1608), and the Mpumalanga Provincial Research and Ethics Committee. Data from the HAALSI study are publicly available at https://dataverse.harvard.edu/dataverse/haalsi.

